# Estimating the real-time case fatality rate of COVID-19 using Poisson mixtures model

**DOI:** 10.1101/2020.04.11.20062190

**Authors:** Paul H. Lee

## Abstract

We proposed using Poisson mixtures model that utilized data of deaths, recoveries, and total confirmed cases in each day since the outbreak. We demonstrated that our CFR estimates for Hubei Province and other parts of China were superior to the simple CFR estimators in the early stage of COVID-19 outbreak.

## INTRODUCTION

The case fatality rate (CFR) of a disease refers to the proportion of death within those infected from the disease. The CFR of an emerging infectious disease can only be determined after its outbreak, but the real-time CFR estimation during the disease outbreak, especially in the early stage, has to be made for public health decisions. There are two naive, simple methods to estimate CFR at a time point *t* using the cumulative number of deaths, recoveries, and confirmed cases at *t*. Denote D(*t*), R(*t*), and C(*t*) be the observed number of deaths, recoveries, and confirmed cases at day *t*. The first simple estimator equals D(*t*)/C(*t*) and the second simple estimator equals D(*t*)/[D(*t*)+R(*t*)]. Note that these two estimators are equivalent after the outbreak when D(*t*)+R(*t*)=C(*t*) [1].

The second estimator usually outperforms the first estimator as there existed a time lag between the diagnosis of disease and death [2-5]. The second estimator at any stage of the outbreak would be unbiased if the proportion of confirmed deaths equals the proportion of unconfirmed deaths [1]. This is the case for severe acute respiratory syndrome (SARS) in 2003 where the time from confirmed diagnosis to death equaled that from confirmed diagnosis to recovery (=23 days [1]). For novel coronavirus disease 2019 (COVID-19), the duration from diagnosis to death was much shorter, with a mean duration of 8-14 days [2-5].

With survivors being hospitalized longer than the deaths, we can expect that the second CFR estimator (hereby referred to as the simple CFR estimator) would overestimate the actual CFR as deaths were more likely to be observed earlier in the outbreak. During the COVID-19 outbreak, many researchers have developed new methods to estimate CFR that replace the nominator or denominator by the relevant figures 8 to 14 days ago to reduce the biasedness of the simple CFR estimator [2-5]. However, this approach ignored the underlying process of how these numbers (deaths, recoveries, and total confirmed cases) were generated and these methods were regarded as invalid [6].

In fact, a statistical modeling approach was developed 15 years ago using the SARS data [1]. The rationale behind this approach is that deaths and recovery conditional at any time point should both follow a parametric distribution (gamma distribution was used to fit the SARS CFR data). The limitation of this approach lies in the estimation of the parameters of this distribution, which require the often-non-public data of time from confirmed diagnosis to death and time from confirmed diagnosis to recovery. Here, we suggest replacing the gamma distribution by Poisson distribution, as we can see below that the knowledge of time to death and time to recovery is not necessary. The proposed model requires data of deaths, recoveries, and total confirmed cases recorded in each day since the outbreak of a population. These data for COVID-19 are publicly-available [7].

Some researchers proposed that the denominator of CFR should also include both those who are diagnosed with the disease and those who are asymptomatic and never diagnosed. In this current study, the CFR for detected disease (also referred to as the symptomatic CFR, or sCFR) was modeled, as the actual number of asymptomatic cases was unknown and our proposed method could not be evaluated.

## METHODS

We assumed that a confirmed case will either die or recover and it is determined *a priori* through a Bernoulli process with a parameter *p*, which is the CFR. If a case is deemed to die, the time to death (in days) follows a Poisson distribution with a mean of λ_*d*_, and the time to recovery for a case who will survive follows a Poisson distribution with a mean of λ_*r*_. We call this model the Poisson mixtures model.

Under this model, the expected number of deaths at day *t*+*a* resulting from the cases confirmed at a particular day *t* should be CFR×C(*t*)×Poi(*a*, λ_*d*_), where Poi(*x*,λ) is the probability mass function of the Poisson distribution with the parameter λ. Similarly, the expected number of recoveries at day *t*+*a* for the cases confirmed at a particular day *t* should be (1-CFR)×C(*t*)×Poi(*a*, λ_*r*_). Therefore, the expected number of deaths at a particular day *t*, D’(*t*), would be Σ_k=0_ ^(*t*-1)^ CFR×C(*t-k*)×Poi(*k*, λ_*d*_), and the expected number of deaths at a particular day *t*, R’(*t*), would be Σ_k=0_ ^(*t*-1)^ (1-CFR)×C(*t-k*)×Poi(*k*, λ_*r*_). We estimated the three parameters of this model by minimizing the chi-square goodness-of-fit statistic, i.e., Σ_k=1_ ^*t*^ (D’(*k*)-D(*k*))^2^/D’(*k*) + (R’(*k*)-R(*k*))^2^/R’(*k*).

The 95% confidence interval of the CFR estimates can be estimated using bootstrap method. Both parametric and non-parametric bootstrap can be used, and here we used non-parametric bootstrap that simulated 1000 datasets with D(*t*), R(*t*), and C(*t*) follow multinomial distribution with the observed proportions as the parameters, and the 2.5^th^ percentile and 97.5^th^ percentile of the estimated CFR from these 1000 datasets were the 95% confidence interval of the corresponding CFR estimate.

The COVID-19 data in Hubei Province, China, and other parts of China up to 10^th^ April 2020 were obtained from the Center for Systems Science and Engineering of John Hopkins University,[7] and all statistical analysis was performed in R 3.6.1. The syntax for our proposed method was available as supplementary material.

## RESULTS

Figure 1 shows the simple CFR estimates and the estimates obtained using our proposed model by time, for Hubei Province and other parts of China, respectively. The 95% CIs of the CFR estimates were shaded in gray. Our proposed CRF estimates for Hubei Province became stable on 5^th^ February, and that for the other parts of China were all very close to the actual CFR on the first day of CFR estimation (25^th^ January). The performance of the Poisson mixtures model for Hubei Province started to overestimate the CFR starting from mid-March when the daily confirmed cases dropped to single-digit onwards and the simple CFR estimator began to converge. Similarly, the performance of our model for other parts of China became inferior to the simple CFR estimator starting from late February when the daily confirmed cases dropped from 200-800 to less than 30.

**Figure 1.**
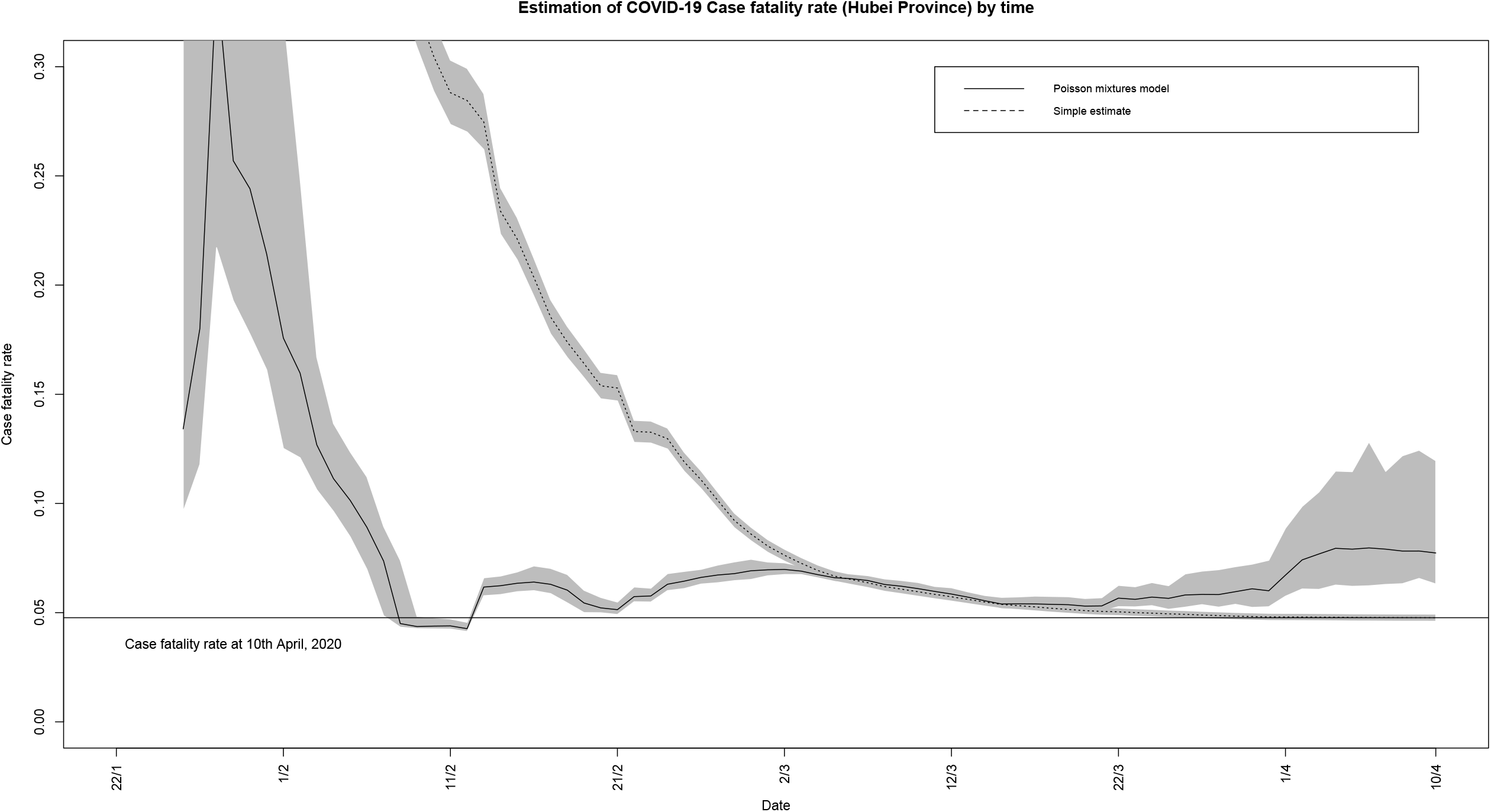
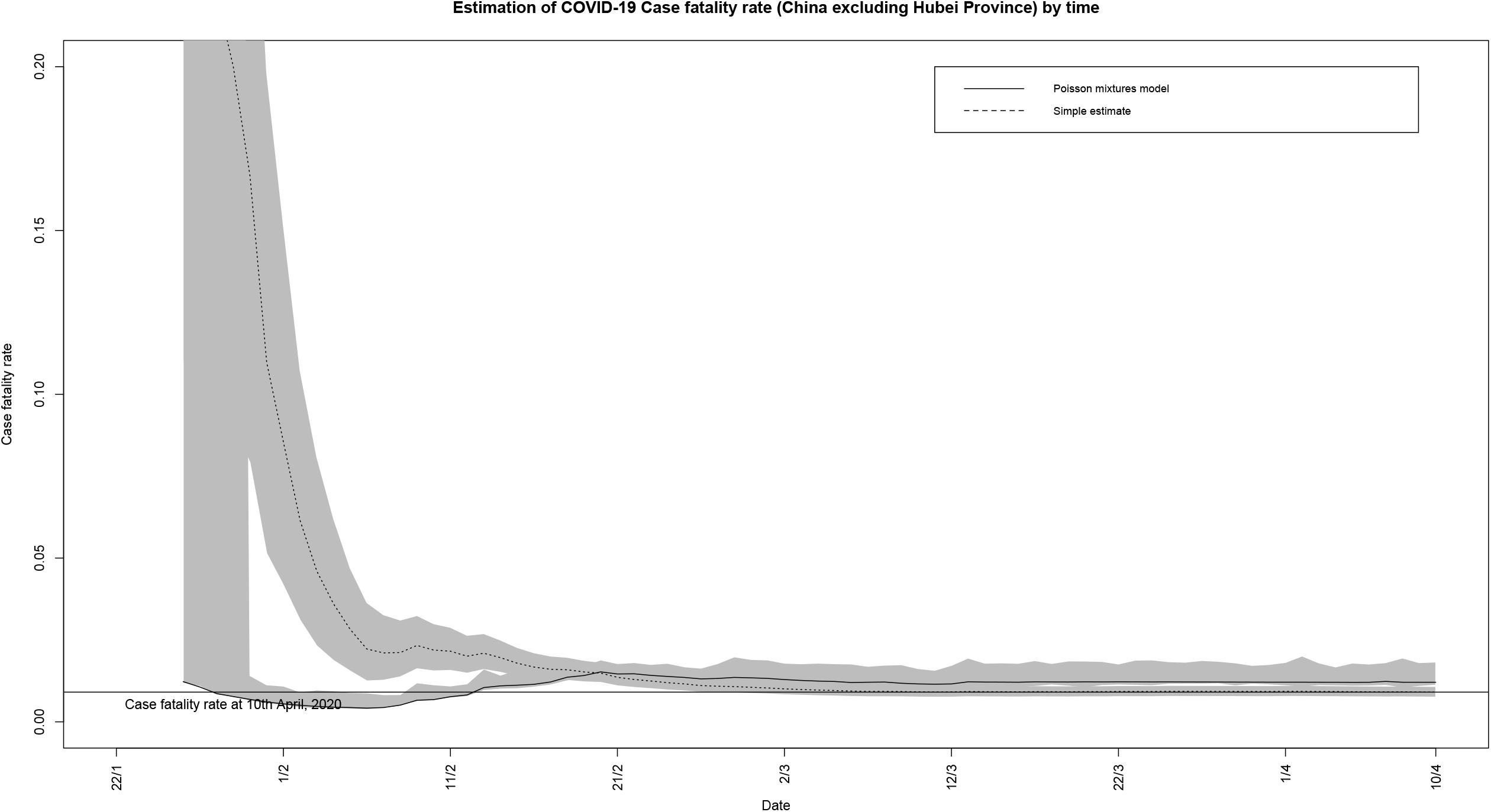
(A) Estimation of COVID-19 Case fatality rate (Hubei Province) by time; (B) Estimation of COVID-19 Case fatality rate (China excluding Hubei Province) by time

## DISCUSSION

Our CFR estimates for Hubei Province and other parts of China were superior to the simple CFR estimators. Making use of all real-time information available for COVID-19, including the number of deaths, recoveries, and total confirmed cases for every single day during the outbreak, our proposed CFR estimates for Hubei Province were acceptable even at the very early stage of the COVID-19 pandemic of mid-February (4.5% to 7%) at which the simple CFR estimators overestimated the actual CFR by more than five folds. Similarly, we achieved CFR estimates for other parts of China at a reasonable level of 1.5% to 1.6% with data up to late January at which the simple CFRs were in the range between 15% and 35%.

Similar to the gamma mixtures model, our proposed model that substituted gamma distribution with Poisson distribution, the CFR estimate was reasonable at the early stage of the disease outbreak, however, the estimate was positively biased in the later stage. We believed that this finding reflected that the actual CFR was changing over time at the later stage of the COVID-19 pandemic, and fitting the data with a constant CFR model may not be appropriate. To test this hypothesis, we can allow the Poisson mixtures model to have one CFR at a time range and another CFR in the other time range, and test the equivalence of these two CFRs. This test of time-varying CFR could also be used to examine the effectiveness of population-level interventions.

The Poisson mixtures model proposed here can be applied to countries in the early and middle stages of the COVID-19 outbreak, for example, the US, Spain, Italy, France, and Germany. However, note that the validity of our model relied on several assumptions, including the constant nature of CFR and the identification of deaths, recoveries, and confirmed cases. The constant CFR assumption may not be appropriate if effective treatments were invented within the disease outbreak.

## Data Availability

The data are publicly available.

https://arcg.is/0fHmTX

